# Evaluating computational assays of chronic fatigue using UK Biobank data

**DOI:** 10.64898/2026.08.21.26360925

**Authors:** Inês Pereira, Herman Galioulline, Ana Grosu, Stefan Frässle, Jakob Heinzle, Zina-Mary Manjaly, Klaas Enno Stephan

**Affiliations:** Translational Neuromodeling Unit (TNU), Institute for Biomedical Engineering, University of Zurich & ETH Zurich, Zurich, Switzerland; Department of Neurology, Schulthess Clinic, Zürich, Switzerland; Department of Health Sciences and Technology (DHEST), ETH Zurich, Zurich, Switzerland; Max Planck Institute for Metabolism Research, Cologne, Germany

**Keywords:** chronic fatigue, computational assay, machine learning, functional magnetic resonance imaging, functional connectivity, effective connectivity, sleep

## Abstract

Chronic fatigue, characterized by persistent physical and/or mental exhaustion, is a frequent and debilitating symptom in medicine. Despite its impact, clinical management remains a challenge. A key problem is the absence of any biomarkers; as a consequence, diagnosis rests entirely on patients’ self-report. This contributes to patient stigmatization and highlights the need for objective diagnostic tools.

In this study, we explored the feasibility of constructing computational assays of chronic fatigue, using clinical and functional neuroimaging data from over 2,200 participants in the UK Biobank. Whole-brain analyses of functional and effective connectivity were followed by machine learning, based on a preregistered analysis plan and a strict separation of training data and held-out test data.

We found that clinical data, including prior medical diagnoses, cancer history, sleep-related information, and alcohol consumption, enabled a statistically significant prediction of chronic fatigue (61% balanced accuracy, p=0.001). Combining clinical information with brain connectivity data again enabled statistically significant predictions (up to 64% balanced accuracy) but did not consistently outperform the model trained on clinical data only. Across all models, sleep-related information, especially insomnia symptoms, emerged as a particularly important feature for prediction.

Our results suggest a high degree of heterogeneity amongst individuals with chronic fatigue. While the predictive performance achieved in this study is not yet sufficient for clinical application, our findings provide a foundation for future developments of objective assays of fatigue. In particular, our results highlight the importance of sleep-related information and suggest new avenues for harnessing neuroimaging information for the prediction of fatigue.

## Introduction

Fatigue is one of the most common symptoms across disciplines in clinical medicine (Bower, 2014; Ceban et al., 2022; Chaudhuri & Behan, 2004; Davies et al., 2021; Gelfand, 2018; Manjaly et al., 2019; Penner & Paul, 2017). Characterized by the experience of extreme and persistent mental and/or physical exhaustion, fatigue can be profoundly debilitating, often impairing activities of daily living and overall quality of life. Yet, despite its impact, clinical management remains a challenge, even at the level of diagnosis: fatigue is a subjective experience, without any biomarker that could corroborate patients’ self-report (Kluger et al., 2013; Manjaly et al., 2019; Penner & Paul, 2017). As a result, patients’ experiences and complaints are open to doubt, leading to ineffective care, repeated medical consultations, and stigmatization. Furthermore, the diversity of self-report questionnaires, with different sources of bias, and absence of objective assays of fatigue pose a significant challenge for clinical trials on fatigue treatments (Byrne, 2022; Fennell et al., 2021; Lakin et al., 2021; Lu et al., 2025). Developing an objective assay for chronic fatigue is therefore an important clinical goal.

Numerous other studies have attempted to predict individuals levels of fatigue from clinical, behavioral, physiological and laboratory data (Adão Martins et al., 2021; Bafna et al., 2021; Baykaner et al., 2015; Cos et al., 2023, 2023; Dias et al., 2024; Du et al., 2023; K. Huang et al., 2025; S. Huang et al., 2018; Hwang et al., 2003; Jiang et al., 2021; Kober et al., 2021; Liu et al., 2026; Luo et al., 2020; Mun & Geng, 2019; Pinto-Bernal et al., 2021; Yao et al., 2021; Zeng et al., 2020; Zuñiga et al., 2020). Unfortunately, there is semantic confusion: many of these studies did not actually predict fatigue (subjective experience), but fatigability (observable decrease in performance during cognitive or motor tasks). Importantly, fatigue and fatigability are two different constructs which are only moderately correlated (for meta-analysis, see Loy et al., 2017). Furthermore, there have been very few attempts to utilize neuroimaging data – particularly functional data – for predicting chronic fatigue. Previous neuroimaging studies were limited by small sample sizes and mostly concentrated on specific disorders in which fatigue is a symptom(Chou et al., 2016; Goñi et al., 2022; Provenzano et al., 2020). At present, no method for predicting fatigue objectively, in a disorder-independent fashion, with sufficient accuracy and low burden for the patient has been established.

Here, we investigated whether this challenge could be addressed by applying machine learning to functional magnetic resonance imaging (fMRI) data during unconstrained cognition (“resting state”), which can be acquired with low burden for patients. Specifically, we sought to exploit recent advances in network approaches that allow for mapping directed interactions (effective connectivity) across the whole brain (Frässle et al., 2017, 2021). In addition to “resting state” fMRI data, we used a broad range of clinical data from the UK Biobank (UKB) (Littlejohns et al., 2020) as features for predicting chronic fatigue in more than 2,200 volunteers. We tested out-of-sample prediction accuracy by splitting the data into training (80%) and held-out test (20%) sets. Before prediction accuracy on the test set was evaluated, models were trained and optimized on the training data according to preregistered analysis plans.

## Methods

The analysis of the data was conducted based on two pre-registered analysis plans. The first analysis plan (https://doi.org/10.5281/zenodo.7835703) details the procedure of participant selection from UKB, whereas the second (https://doi.org/10.5281/zenodo.8185698) defines the statistical and machine learning analyses. Any deviation from these analysis plans is indicated in this manuscript.

### Selection of participants

Participants for this study consisted of subjects who fulfilled the following two conditions (Figure 1 and 2):

1. They had completed the online pain questionnaire (OPQ) (*Pain Web Questionnaire, Version 2.1*, 2022), in particular, the following questions from Section J, on fatigue:

○ Do you have persistent or recurrent tiredness, weariness or fatigue that has lasted for at least 6 months? (UKB data field 120114)
○ The Fatigue Severity Scale (FSS) questions (UKB data fields 120119 to 120127)
2. They had had an MRI session in the 6 months preceding the completion of the OPQ. During the imaging assessment, resting-state functional MRI (rs-fMRI) images were collected, as well as clinical information, via a touchscreen questionnaire and a subsequent verbal interview.

**Figure 1:**
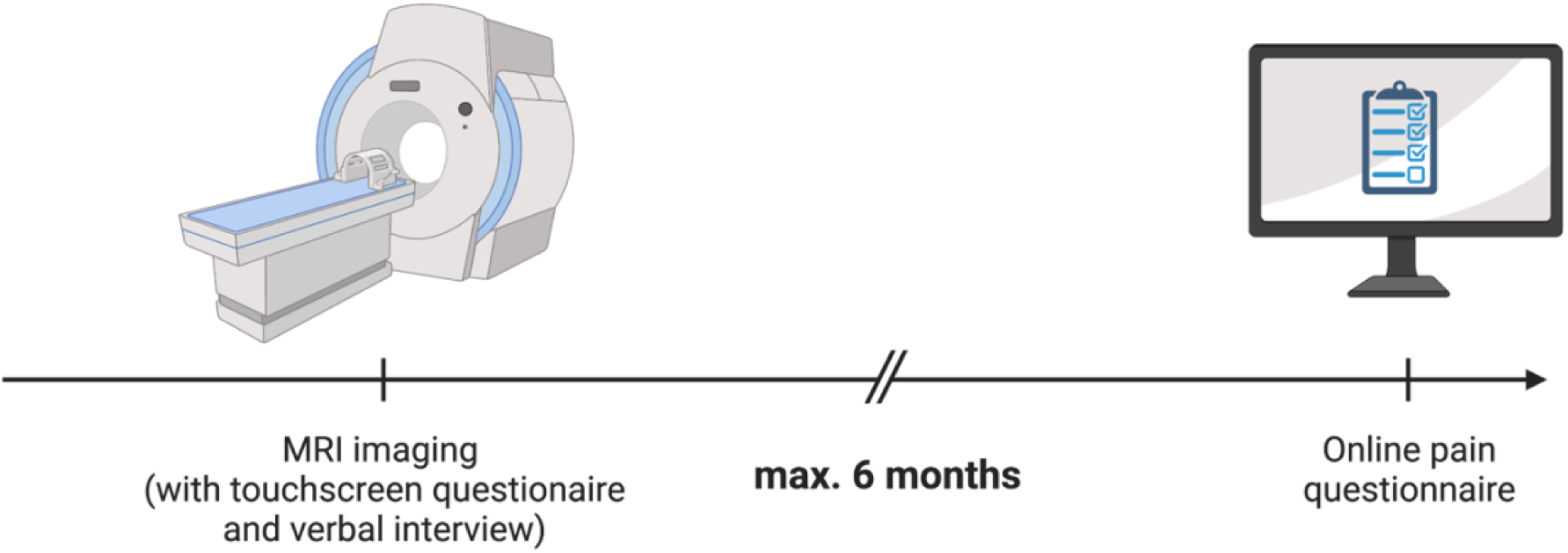
Base criteria for participant selection. Briefly, we selected participants who completed Section J (on fatigue) of the online pain questionnaire (OPQ) and, in the preceding 6 months, attended an imaging assessment. During the imaging assessment, resting-state functional MRI images were collected, as well as clinical information, via a touchscreen questionnaire and a subsequent verbal interview. Figure created in https://BioRender.com

**Figure 2:**
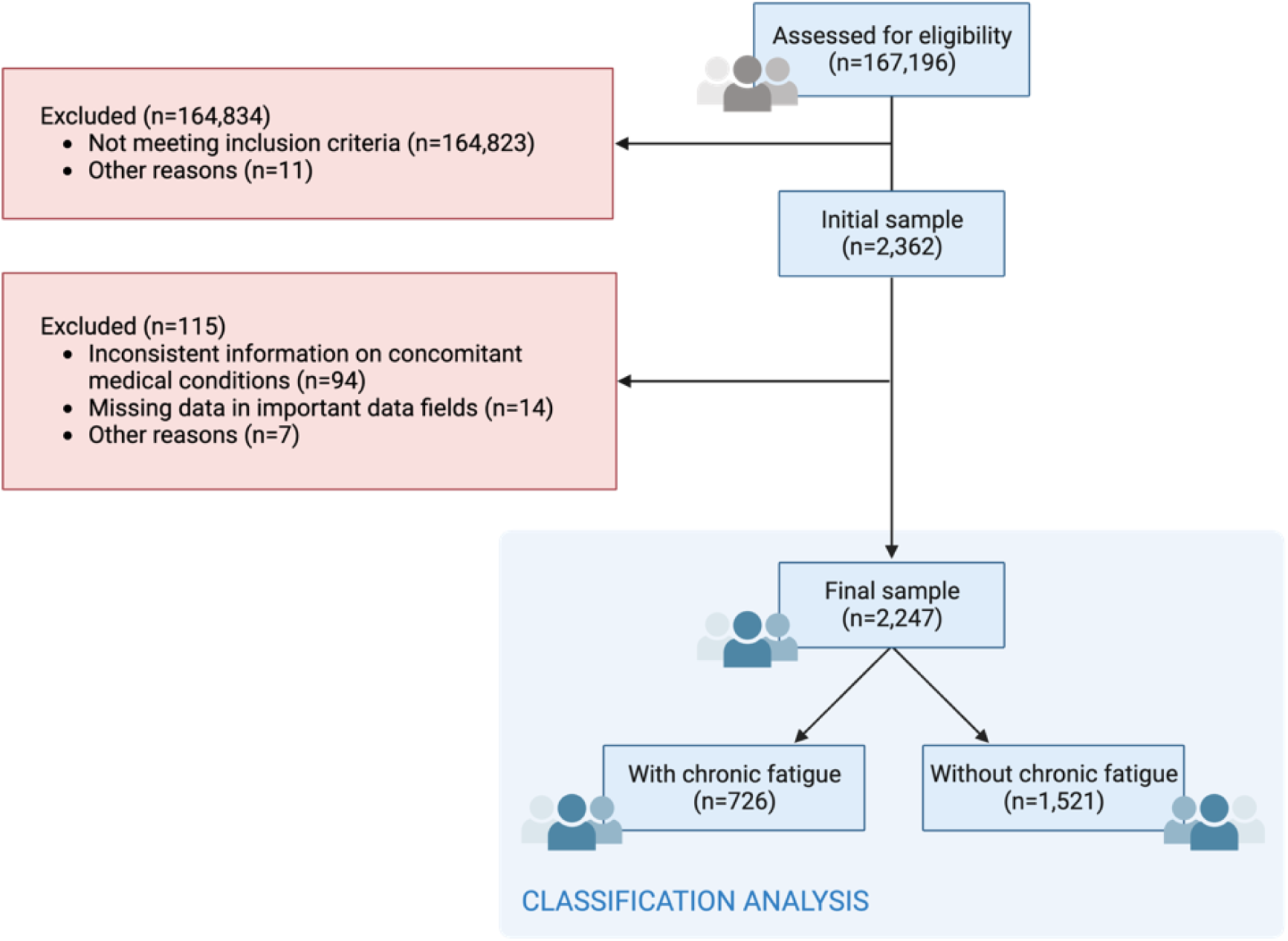
CONSORT flow diagram. 167,196 individuals that had completed the online pain questionnaire (OPQ) were assessed for eligibility. Of these, 2,362 individuals had had an MRI session in the 6 months preceding the completion of the OPQ. 11 subjects withdrew from the UK Biobank; thus, their data were excluded from analyses. From the 2,362 meeting the initial inclusion criteria, a further 115 were excluded for the following reasons: (1) inconsistent information regarding the presence of concomitant medical conditions (when comparing the data from the touchscreen questionnaire and medical interview, both done at imaging), (2) missing data in several features of interest (either because of NaNs or because of answers such as “do not know” or “prefer not to respond”). The classification dependent variable was defined as the binary answer to the question in data field 120114 (Do you have persistent or recurrent tiredness, weariness or fatigue that has lasted for at least 6 months?). 1521 (i.e., two thirds of the) subjects answered “No”, whereas 726 (one third) reported that they did, indeed, suffer from these symptoms. Figure created in https://BioRender.com

167,196 individuals that had completed the OPQ were assessed for eligibility. 2,362 subjects met the above criteria; of these, 115 were excluded due to missing data or data inconsistencies (see Figure 2).

### Clinical features

#### Dependent variable

The dependent variable was defined as the binary answer to the question in UKB data field 120114: *Do you have persistent or recurrent tiredness, weariness or fatigue that has lasted for at least 6 months?* 726 participants (approx. one third) responded with “Yes”.

#### Independent variables

The imaging visit included a phased clinical assessment. Participants were first asked to provide information about their clinical history via a touchscreen questionnaire. A verbal interview was subsequently conducted with a UKB staff member.

Using both sources, an individual’s medical history was captured comprehensively by including information about 445 diagnoses. We started by defining a one-hot encoding for all selectable diagnoses from UKB data field 20002 (https://biobank.ctsu.ox.ac.uk/ukb/field.cgi?id=20002), as defined by UKB data-coding 6 (https://biobank.ndph.ox.ac.uk/showcase/coding.cgi?id=6). Where possible, information from the touchscreen questionnaire of the imaging session was also included. For a given medical condition, the feature was set to “true” if the participant reported the diagnosis in either the touchscreen questionnaire or the verbal interview. Time since diagnosis was also encoded, adding another 445 features. However, the feature set resulting from this initial selection procedure was sparse and strongly affected by multicollinearity. Therefore, the encoding of the clinical features was simplified. Firstly, diagnoses were grouped into 11 top-level categories, e.g. cardiovascular, gastrointestinal, or endocrine disorders, as defined by the UKB classification (https://biobank.ctsu.ox.ac.uk/ukb/field.cgi?id=20002). For each, a new time-since-diagnosis feature was created, summing the time since diagnosis (in years) over the several diagnoses under that top node, for each subject.

Beyond information about general medical history, we included additional details about cancer history, sleep, and alcohol intake. Cancer history, assessed via UKB data fields 2453 and 20001, was coded as a binary feature (true if cancer was reported in either field). Sleep was assessed via questions on: (1) average sleep duration (UKB data field 1160), (2) the frequency of insomnia (UKB data field 1200) and (3) the tendency for daytime dozing/sleeping (UKB data field 1220). Alcohol intake was represented by UKB data field 1558. For the latter two sleep data fields and alcohol intake, we defined a one-hot encoding to cover all possible answers.

Overall, the feature engineering process described above resulted in the definition of 27 clinical features.

### Brain imaging features

We used the preprocessed rs-fMRI data provided by the UKB. For details, please see http://biobank.ndph.ox.ac.uk/ukb/ukb/docs/brain_mri.pdf and Alfaro-Almagro et al. (2018). Briefly, the data were collected in three imaging centers using identical 3T Siemens Skyra with 32-channel receive head coils. The rs-fMRI acquisition lasted 6 minutes (490 timepoints, TR=0.735 s), with 2.4 mm isotropic resolution and 8× multislice acceleration. The UKB’s preprocessing pipeline included group spatial ICA decomposition. The resulting components were classified by UKB as signal/noise (for a list of these components, please see https://www.fmrib.ox.ac.uk/ukbiobank/) and then fed into dual regression to give a set of subject-level timeseries, at dimensionalities of either 25 or 100. From these, 21 and 55 components were labeled as non-artefactual, respectively. In the following, we refer to these as the ICA21 and ICA55 datasets.

Functional connectivity matrices were computed using Pearson’s correlation coefficient. To capture directed interactions, effective connectivity was estimated using regression dynamic causal modeling (rDCM), a computationally efficient DCM variant for fMRI data (Frässle et al., 2017, 2021). We used the original MATLAB implementation of rDCM within the TAPAS software suite: https://www.translationalneuromodeling.org/tapas.

The number of connectivity features generated thus depended on the timeseries used and on the connectivity method employed:

- Functional connectivity (ICA21): 210 features
- Functional connectivity (ICA55): 1485 features
- Effective connectivity (ICA21): 441 features
- Effective connectivity (ICA55): 3025 features

### Machine learning analyses

The machine learning pipeline was built using the Python package scikit-learn (https://scikit-learn.org/). We employed logistic regression with elastic net regularization (Zou & Hastie, 2005) and nested cross-validation. Cost-sensitive classification was performed by setting the class weights hyperparameter to ‘balanced’ (for details, see the scikit-learn documentation: https://scikit-learn.org/stable/modules/generated/sklearn.linear_model.SGDClassifier.html). Model performance was evaluated using balanced accuracy (BA), and the statistical significance of predictions was assessed via permutation testing (1,000 permutations).

### Nested cross-validation: choosing a winning model

During nested cross-validation (applied to the training data), each outer loop generates one winning model. Thus, across loops, one is often faced with several winning models, with different hyperparameter values. Following our preregistered analysis plan, we selected the final model (to deploy on the test set) via majority voting. In case of a tie, we reran one round of 10-fold cross-validation on the full training set with the final candidate models (those involved in the tie), selecting the best-performing model. This final model was then retrained on the entire training set (i.e., estimated the model parameters, with the hyperparameters now fixed) before evaluation on the test set. See Supplementary Material for a visual overview.

### Feature importance

We assessed feature contributions using logistic regression weights and SHAP (SHapley Additive exPlanations) values (Lundberg & Lee, 2017). Based on the Shapley values from Game Theory (Shapley, 2016), this method generalizes this approach to machine learning, indicating how much each feature contributed to the overall prediction. We used the Python implementation by the original authors (https://github.com/shap/shap), with a KernelExplainer, on the test set. SHAP values provide explanations for individual features and observations (subjects) and indicate the directionality of a feature’s influence on the prediction. Comparing SHAP results and logistic regression weights, we verified that the feature importance results were concordant.

### Training and test sets – Rules of access

As mentioned previously, we implemented an 80/20 split of the data into training and test sets, with randomly chosen 20% of study participants allocated to the test set. Once the analysis pipeline was finished on the training set, the final step was to test the predictive accuracy of a given model on the held-out test data. However, before this was done, an independent code review of the analysis code was conducted (Pereira, Galioulline et al., 2026) with implementation of any necessary corrections and adjustments.

### Deviations from the analysis plan

We had pre-registered two analyses: a classification analysis (reported here) and a regression analysis with the goal of predicting FSS scores derived from the OPQ. However, the sample available for the regression analysis consisted of only 91 UKB participants who met the inclusion criteria and had data of sufficient quality. Given this small sample size, even on the training set, predictions proved to challenging, despite elastic net regularization. Therefore, we did not pursue this analysis further. Furthermore, we conducted several post-hoc analyses to understand the importance of a sleep-related feature for predictions, as well as that of clinical versus imaging features; see Results for details.

## Results

### Prespecified analyses

Given the above-described feature engineering procedure, we constructed four combined datasets. Each contained the same clinical, but different connectivity features (functional or effective connectivity, from the ICA21 or ICA55 timeseries). Our analyses showed that all datasets enabled significant predictions on both training and test data (see Table 1 for results on the test set). The BA across all four datasets ranged from 60.0% to 63.5%.

**Table 1:** Results of permutation testing on the test set. The models tested here were trained either on the combined datasets (i.e., datasets containing both clinical and imaging data) or on the clinical data only. func21 and func55 correspond to the datasets containing the clinical features, as well as the functional connectivity features from the ICA21 and ICA55 timeseries, respectively. The rdcm21 and rdcm55 datasets contain the rDCM (effective) connectivity estimates from the ICA21 and ICA55 timeseries, respectively. BA = balanced accuracy. Uncorrected p-values are displayed. In bold are those that survive Bonferroni correction for multiple comparisons (threshold for significance after Bonferroni correction is 0.005).

| Dataset | Feature set | BA on test set<br>(p-value) | Difference in BA compared to using<br>only ‘3_insomnia’<br>(p-value) |
| --- | --- | --- | --- |
| **func21** | Clinical features + functional connectivity features from the ICA21 timeseries | 63.5%<br>**(p = 0.001)** | 0.0620<br>**(p = 0.002)** |
| **rdcm21** | Clinical features + effective connectivity features from the ICA21 timeseries | 60.5%<br>**(p = 0.001)** | 0.0319<br>(p = 0.066) |
| **func55** | Clinical features + functional connectivity features from the ICA55 timeseries | 60.4%<br>**(p = 0.001)** | 0.0302<br>(p = 0.133) |
| **rdcm55** | Clinical features + effective connectivity features from the ICA55 timeseries | 60.0%<br>**(p = 0.001)** | 0.0269<br>(p = 0.152) |
| **clinical** | Clinical features | 61.2%<br>**(p = 0.001)** | 0.0386<br>(p = 0.050) |

Analysis of the associated logistic regression weights and SHAP values showed that the sleep-related feature ‘3_insomnia’ played a prominent role in prediction in all tested models (for details, see Supplementary Material). Feature ‘3_insomnia’ corresponded to the binary encoding of the answer “usually” to the question “*Do you have trouble falling asleep at night or do you wake up in the middle of the night?*” (UKB data field 1200). To better understand the importance of this predictor, as well as that of clinical versus imaging features, we conducted several post-hoc analyses that were not described in our analysis plans.

### Post-hoc analyses

We first trained a new model using only the clinical features and no imaging-related predictor. With a BA of 61.2%, this model was marginally better than most models using combined (clinical plus imaging) data, with the exception of the model trained on clinical data and functional connectivity features from the ICA21 timeseries (Table 1). Next, we tested a model containing feature ‘3_insomnia’, encoded as a binary feature, as the sole predictor. This achieved a BA of 57.3% on the test data. While numerically lower, this accuracy was statistically not significantly different from that obtained by most models using clinical or clinical+neuroimaging datasets, with the exception of the func21 dataset (see Table 1).

## Discussion

Our key findings are as follows: Clinical features, including prior medical diagnoses, cancer history, sleep-related information, and alcohol consumption, enabled a statistically significant prediction, with a BA of 61.2% on the test set. Combining this clinical information with neuroimaging (functional and effective connectivity estimated from rs-fMRI data), we achieved a balanced accuracy ranging from 60.0% to 63.5%, with the best model combining clinical features with functional connectivity estimates from the ICA21 timeseries. Surprisingly, only the latter model outperformed the model trained on clinical data only. Finally, we noted the particularly important role of a specific feature, the experience of insomnia, for prediction. A model using only this feature achieved a BA of 57.3% on the test set. While all other, more complex models display higher BA scores, the difference was only statistically significant when using the func21 dataset. Thus, our results suggest that the main driver of prediction is sleep-related information, with clinical and connectivity features improving results, but not consistently in a statistically significant manner.

Sleep is in fact recognized as a secondary cause of fatigue, e.g. in specific patient populations such as multiple sclerosis (Attarian et al., 2004; Foschi et al., 2019; Rouault et al., 2023; Veauthier & Paul, 2014). However, we caution against interpreting our finding as chronic fatigue simply corresponding to chronic tiredness due to insomnia. First, normal sleep time does not necessarily provide relief for chronic fatigue (cf. diagnostic criteria for chronic fatigue syndrome/myalgic encephalomyelitis; Gluckman & Grach, 2026). Even more importantly, sleep disturbances have profound impact on homeostatic and allostatic regulation (McEwen, 2006), a topic at the core of mechanistic explanations of chronic fatigue (Stephan et al. 2016; Manjaly et al. 2019). Given that we are dealing with a heterogeneous sample from the general population, it is possible that sleep-induced disruptions of homeostasis might constitute a “common factor” that explains the presence of fatigue moderately well in a majority of cases.

Our study has strengths and limitations. Strengths include pre-registered analysis plans, multi-modal datasets, and a large sample size (N>2,200). In addition, we conducted a strict separation of training and test data, with no leakage of information, protecting against overfitting. This aspect was also explicitly checked during the independent code review (Pereira, Galioulline et al., 2026). Finally, we used advanced methods for computing effective (directed) connectivity estimates across the whole brain that have proven useful for other clinical predictions (Galioulline et al., 2023).

Turning to the limitations of our study, UKB offers limited information for detecting individuals suffering from chronic fatigue. The main source is a single entry (data field 120114), as described above. While practical and complying with the duration used to define chronic fatigue (6 months), relying on one data field for a concept as complex as fatigue is not ideal. Additionally, the OPQ in UKB includes questions which essentially reproduce the FSS, an established fatigue questionnaire by Krupp et al., (1989). However, these questions only refer to the previous week and thus do not capture chronic fatigue well. Additionally, only a small sample of UKB participants could be used for our planned regression-based prediction of FSS scores, preventing us from pursuing it further, as mentioned above.

Another limitation concerns the fMRI measurements available in UKB. For investigating fatigue, rs-fMRI may be too unspecific; additionally, the relatively short duration of the measurements (6 minutes) is not ideal for reliability (Birn et al., 2013). Furthermore, the standard preprocessing procedure of the UKB fMRI data means that subcortical areas, thought to be of major relevance for the experience of fatigue (Stephan et al., 2016), have very little representation in the extracted timeseries.

Notwithstanding these limitations, our study makes several important contributions. First, we show the feasibility of predicting symptoms of chronic fatigue out-of-sample, using only clinical history and sleep data and within a much larger and more heterogeneous sample than previous attempts (e.g. Rouault et al., 2023). While the predictive accuracy we achieved is not yet sufficient for clinical application, our findings provide a foundation for future developments of objective assays of fatigue. In particular, our results highlight the utility of sleep-related information and suggest new avenues for harnessing neuroimaging information for the prediction of fatigue. With regard to sleep, using more quantitative and objective assessments (e.g. using wearables) than in this study might further enhance the predictive utility of sleep data. Concerning fMRI, it may be important to obtain measurements under more constrained cognitive contexts that are of direct relevance for the experience of fatigue and ensure coverage of subcortical regions including the brainstem. We hope that extensions along these lines will ultimately help establish objective assays of fatigue.

## Data Availability

This research was conducted using the UK Biobank Resource under Application Number 60679.

https://www.ukbiobank.ac.uk/

## Data and code availability

Our study uses data provided by the UK Biobank, which is made available to all qualified researchers on application. We provide our code for dataset definition and model training under https://gitlab.ethz.ch/tnu/code/pereiraetal_fatigue_ukb.

## Acknowledgments

KES acknowledges support by the René and Susanne Braginsky Foundation, the ETH Foundation, and the Precision Medicine for Integrative Mental Health Consortium by University Medicine Zurich (UMZH). This research was conducted using the UK Biobank Resource under Application Number 60679.

## Supplementary material

### Capturing information on the causes of fatigue: available data

For this study, we attempted to capture as many of the factors listed previously without adding a prohibitive number of features to the model. To capture information on concomitant medical and neurological or psychiatric conditions, the following data fields from the imaging session were considered:

- [Touchscreen questionnaire] Has a doctor ever told you that you have had any of the following conditions? Heart attack, Angina, Stroke, High blood pressure (Data field 6150)
- [Touchscreen questionnaire] Has a doctor ever told you that you have had any of the following conditions? Blood clot in the legs (DVT), Blood clot in the lungs, Emphysema/chronic bronchitis, Asthma, Hayfever, allergic rhinitis or eczema (Data field 6152)
- [Touchscreen questionnaire] Has a doctor ever told you that you have had any other serious medical conditions or disabilities? (Data field 2473)
- [Touchscreen questionnaire] Has a doctor ever told you that you have had cancer? (Data field 2453)
- [Touchscreen questionnaire] Has a doctor ever told you that you have diabetes? (Data field 2443)
- [Verbal interview] Non-cancer illness code, self-reported (Data field 20002), paired with age/date first diagnosed (Data field 87)
- [Verbal interview] Cancer illness code, self-reported (Data field 20001), paired with age/date first occurrence (Data field 84)

Individual who answered “Yes” in Data field 2473, but then presented no data in data field 20002 were considered inconsistent and removed for the sample of participants. This affected 94 subjects out of the initial sample of 2,362 (Figure 2). Data field 2473 was subsequently not included as a feature for the model, given that it captured no additional information once the inconsistencies in the data had been removed.

To capture information on **sleep and alcohol consumption**, we considered the following data fields from the touchscreen questionnaire at the imaging session:

- [Touchscreen questionnaire] *About how many hours sleep do you get in every 24 hours? (please include naps)* (Data field 1160)
- [Touchscreen questionnaire] *How likely are you to doze off or fall asleep during the daytime when you don’t mean to? (e.g. when working, reading or driving)* (Data field 1220)
- [Touchscreen questionnaire] *Do you have trouble falling asleep at night or do you wake up in the middle of the night?* (Data field 1200)
- [Touchscreen questionnaire] *About how often do you drink alcohol?* (Data field 1558)

7 subjects who reported “Do not know” or “Prefer not to answer” under data field 1160 were excluded from the sample (Figure 2). This was done to facilitate the encoding of this feature and avoid data imputation (see Section: “Clinical features”).

### UK Biobank’s fMRI preprocessing pipeline

The resting-state fMRI (rs-fMRI) data preprocessing pipeline included (1) EPI & gradient distortion correction (GDC) unwarping, (2) motion correction using MCFLIRT, (3) highpass temporal filtering (Gaussian-weighted least-squares straight line fitting, with sigma = 50.0s) and (4) removal of structured artefacts by ICA+FIX processing (Alfaro-Almagro et al., 2018). A transform from T1 space to standard MNI space was used to resample each subject’s preprocessed timeseries dataset into standard space.

In addition, UK Biobank’s preprocessing included a group spatial ICA decomposition, performed using MELODIC. The resulting components were classified by UKB as signal/noise^1^, and then fed into dual regression to give a set of subject-level timeseries, at dimensionalities of either 25 or 100. From the 25-and 100-dimensional timeseries, 21 and 55 components were labeled non-artefactual, respectively (Alfaro-Almagro et al., 2018). These sets of ICA maps can be viewed as “parcellations” of cortical and sub-cortical grey matter, however, ICA maps are not binary masks but contain a continuous range of values. In addition, they can overlap each other, and a given map can include multiple spatially separated regions.

For this analysis, we used the subject-specific ICA timeseries under rfMRI_*.dr/dr_stage1.txt, where * is the original timeseries dimensionality (25 or 100). Our functional and effective connectivity analyses made use of the non-artefactual subset of the provided dual regression timeseries, thus working with timeseries of dimensionality 21 and 55 (referred to henceforth as the ICA21 and ICA55 timeseries).

### Nested cross-validation: choosing a winning model

**Figure A:**
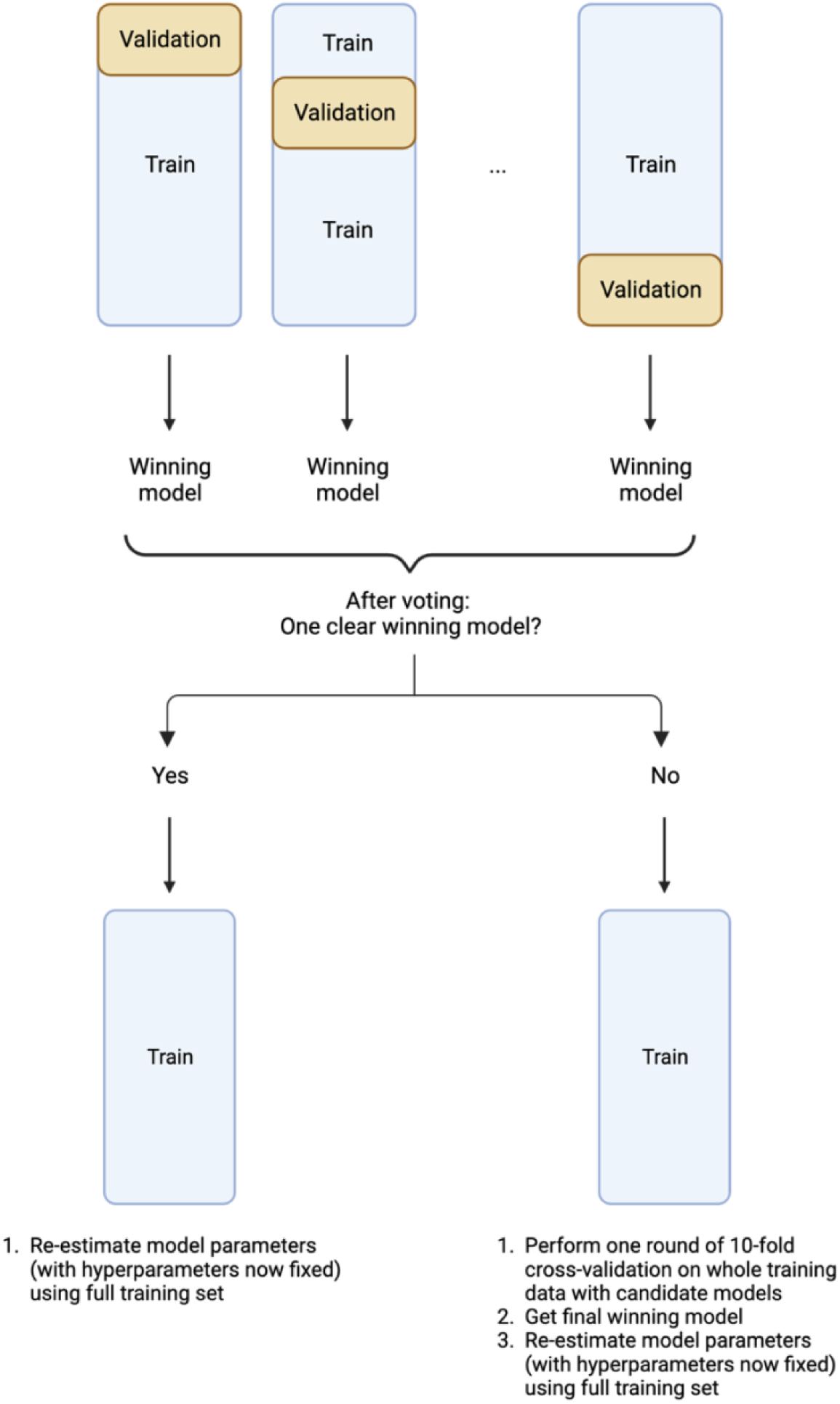
Choosing a winning model during nested cross-validation. If, via majority voting, no clear winning model arose from the nested cross-validation procedure, all candidate models (those involved in a tie) were used to perform a final 10-fold (simple) cross-validation round on the whole training data. This yielded a final winning model. As a last step, we used the entirety of the training data to re-estimate the winning model’s parameters (with the hyperparameters now fixed). Figure created in https://BioRender.com

### Results - Training data

**Figure B:**
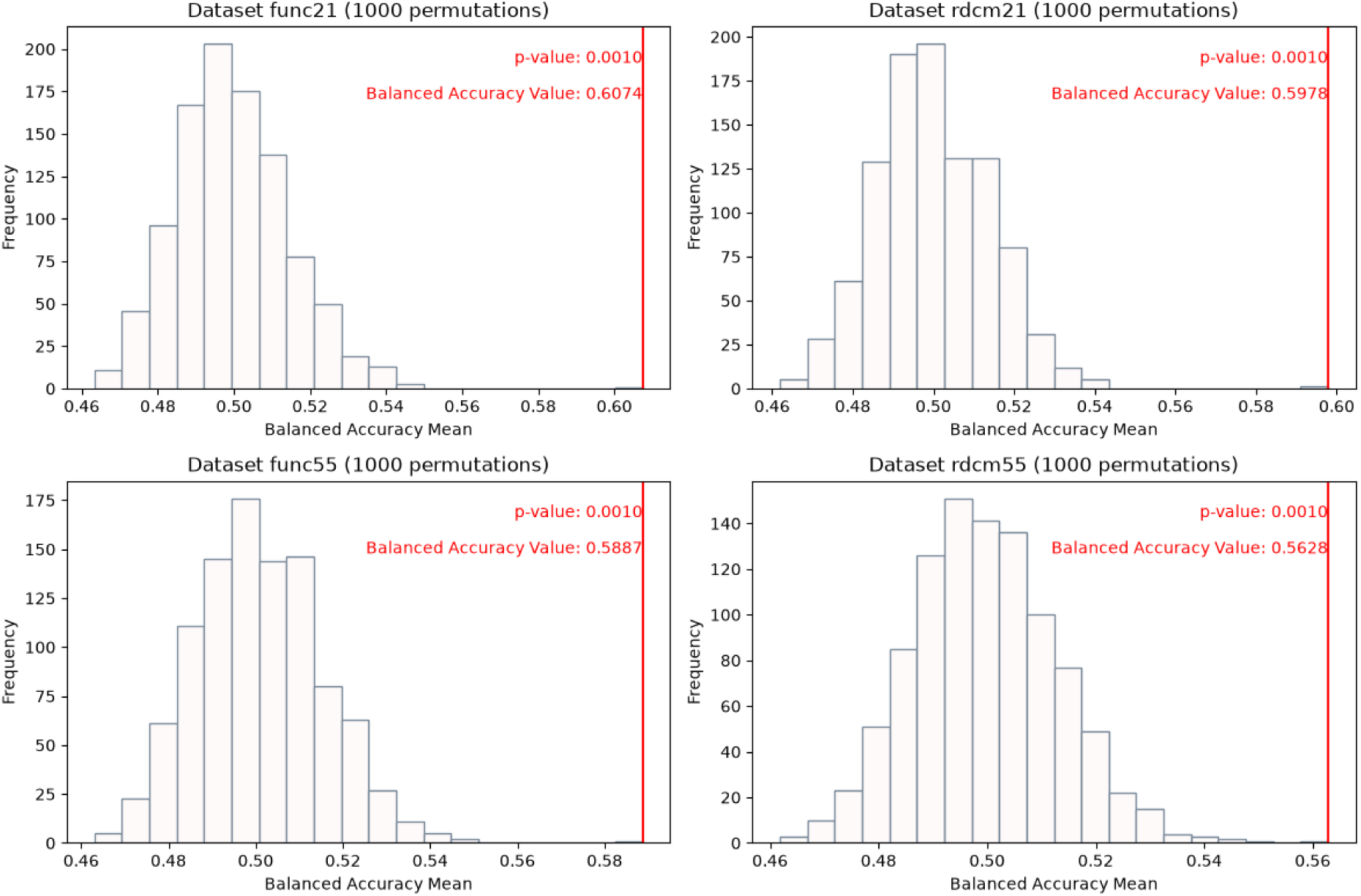
Results of permutation testing on the training data. All models tested here were trained on combined datasets (i.e., on datasets comprising both clinical and imaging data). func21 and func55 correspond to the datasets containing the clinical features, as well as the functional connectivity features from the ICA21 and ICA55 timeseries, respectively. The rdcm21 and rdcm55 datasets contain the rDCM (effective) connectivity estimates from the ICA21 and ICA55 timeseries, respectively. BA = balanced accuracy. Uncorrected p-values are displayed..

### Results - SHAP values

**Figure C:**
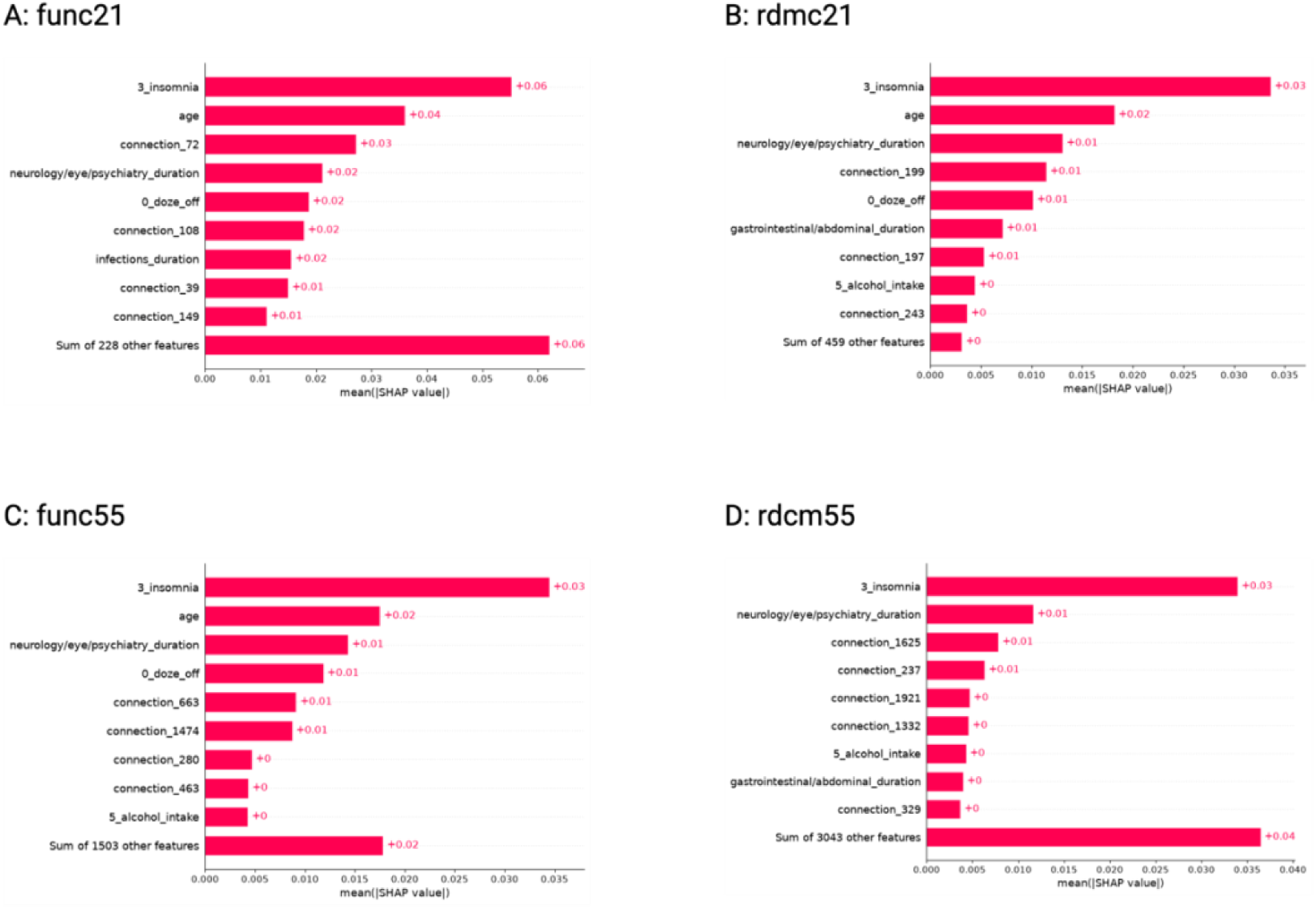
Mean absolute SHAP values of the 10 most important SHAP values for each model using the combined datasets. The func21 and func55 datasets contain the clinical features and functional connectivity features from the ICA21 and ICA55 timeseries, respectively. The rdcm21 and rdcm55 datasets contain the rDCM (effective) connectivity estimates from the ICA21 and ICA55 timeseries, respectively.

## Footnotes

1 Refer to published lists of relevant components here: https://www.fmrib.ox.ac.uk/ukbiobank/

## Notes

### Competing Interest Statement

The authors have declared no competing interest.

### Author Declarations

This research was conducted using the UK Biobank Resource under Application Number 60679.

